# Prevalence of lead poisoning among artisanal gold miners in French Guiana in 2022

**DOI:** 10.1101/2025.01.31.25321474

**Authors:** Maylis Douine, Virgile Korsec, Alice Sanna, Lorraine Plessis, Teddy Bardon, Antoine Adenis, Mathieu Nacher, Martha Suarez-Mutis, Stephen Vreden, Olivier Mathieu, Yann Lambert

## Abstract

**Objectives:** Lead poisoning is a major public health problem worldwide. In French Guiana, a French overseas territory in South America, several studies have highlighted the massive lead impregnation of the local population, but the risk factors have not yet been fully elucidated. People working in informal gold mining share part of their lifestyle with French Guianese communities. The aim of this was therefore to estimate the level of lead poisoning in this population and the factors associated with it.

**Methods:** This cross-sectional descriptive study was based on data collected by questionnaire and blood sampling. Persons working in informal gold mines were enrolled on the logistic rear bases on the Surinamese and Brazilian sites of the bordering rivers. Blood lead levels were measured on dry blood spots (50 µl). Using a threshold of 100 μg.L^−1^, single and multiple regressions were used to assess the associated factors.

**Results:** Among the 526 persons included, the median age was 38 years and 73.5% were men. The prevalence of lead poisoning was 44.7 % (95%CI=40.4%-49.0%). The factors associated with a BLL over 100 μg.L-1 were: amount of time spent in gold mining (OR=1.31 [1.09-1.58], occupation with mud exposure (OR=1.67 [1.13-2.48]), working in the Southwestern region (OR=2.09 [1.34-3.27]) and consuming game (OR=1.58 [1.06-2.36]).

**Conclusion:** The people working on gold mining sites are highly exposed to lead poisoning. The risk factors are suggestive of environmental contamination and differ from those suspected in the population of French Guiana.

**Research in context:** *Evidence before study:* Lead poisoning has a major impact on health. Although, its risk factors are well known in the Western world, they are different in the Amazon. In French Guiana, a French overseas territory in South America, lead poisoning is very common in the local population, but the risk factors are still poorly understood. So far, the suspected sources of lead poisoning are the bioaccumulation of lead in cassava flour, lead contained in hunting ammunition, lead used to weigh the fishing nets, and lead spills from used lead batteries.

*Added value of this study:* This study shows, for the first time, the high lead impregnation of the population living and working on illegal gold mining sites in French Guiana. Although some aspects of their lifestyles are shared with the local population, the risk factors we observed were different from those commonly hypothesized. We found that length of time spent in gold mining, specific geographic areas and activities in close contact with mud were significantly associated with lead poisoning.

*Implications of all the available evidence:* The contrast between the factors associated with lead poisoning in the local population and in the population working at the gold mining sites allows to propose new hypotheses and research questions to be proposed about potential sources of contamination, for example related to local geology.

## Introduction

Lead poisoning is a global public health problem that kills one million people each year. Millions more, many of them children, are exposed to low levels of lead, resulting in lifelong health problems including anemia, hypertension, immunotoxicity and reproductive organs toxicity, and often irreversible neurological and behavioral effects [1,2]. In 2015, the World Health Organization (WHO) selected a concentration of 50 _μ_g.L^−1^ as the threshold required to trigger investigations to identify and reduce lead exposure. However, this definition should not obscure the understanding that there is no threshold effect in childhood lead poisoning [3].

French Guiana (FG), a French overseas territory bordering Brazil and Suriname, and largely covered by rainforest, has a lead poisoning problem [4]. Lead poisoning was first detected in FG in Mana in 2011, in a 3-year-old girl with a blood lead level of 1700 μg.L-1. Investigation revealed a cluster of 35 intoxications > 50 μg.L-1. No source of exposure was identified, but dietary exposure was suspected. [5,6]. Since then, several studies have been conducted to assess the burden of lead poisoning in FG. The prevalence of poisoning in children was enormous, measured at 20.1 % [15.9-24.6] in 2015-2016 [7]. The suspected causes identified by the French Food and Environment Agency (ANSES) included lead bioaccumulated in *couac*, a type of cassava flour consumed in the Guiana Shield, lead in hunting ammunition, lead used to weigh fishing nets, and lead spilled from used lead batteries [8].

The local populations affected by lead poisoning in FG share some aspects of their way of life with another population living nearby, the workers at artisanal and small-scale gold mining (ASGM) sites. There are an estimated 11,000 gold miners, mostly from Brazil, and their mining activities are mostly illegal (7). Their lifestyle exposes them to numerous health problems, such as vector-borne diseases, and their distance from the health system often results in poor access to care, treatment interruptions and poor follow-up, leading to a lack of public health data on this population [9,10]. Because of the partly shared context with the French Guianese population, we hypothesized that the gold miner population may also be affected by lead poisoning through similar sources of poisoning.

The present study therefore assesses whether the population working in ASGM is also affected by lead poisoning. The objectives were to describe the prevalence of lead poisoning and to identify the risk factors in the population of gold mine workers in 2022.

## Material and methods

### Study design

This study was a cross-sectional descriptive survey based on data collected by questionnaire and biological sampling. This study is ancillary to an interventional research project called Curema which aims to evaluate a new strategy to eliminate malaria in this vulnerable population [11]. The inclusions took place from September to December 2022 in the logistical rear bases of FG illegal gold mining using opportunistic sampling. These were located on the Brazilian border of the Oyapock river and on the Surinamese border of the Maroni river.

### Study population

The target population was people working on illegal gold mining sites in FG (9). Inclusion criteria were: presence on a gold mining site in the seven days prior to the inclusion, individual consent, and age over 18 years. The sample size was calculated based on the main objective of this survey, which was to assess malaria prevalence (estimated=25%, alpha=5%, 95%CI, N=281 for each border, thus 562).

### Survey data, blood collection and analysis

The questionnaire included variables on socioeconomic status, gold mining activity, and lifestyle. A rapid and structured clinical examination focused on malaria was performed by a physician. Blood pressure was measured. A venous blood sample was taken from each participant in an EDTA tube. Using a micropipette, 50 µl of total blood was then applied to a filter paper and stored with desiccant in a hermetic box. The measurement of blood lead on these dried blood spots was performed by the toxicology laboratory of the Centre Hospitalier Universitaire de Montpellier using an ICP-MS method (iCAP-Q, ThermoFisher Scientific^TM^, Illkirch, France). Briefly, the whole blood spot was isolated and digested for 30 minutes in 5 mL of 5% ultrapure nitric acid solution. Lead concentration, based on isotope 208, was determined using an 8-point calibration curve (20 to 2,000 µg.L^−1^) generated from a 1 g.L^−1^ standard solution (Merck^TM^, Darmstadt, Germany). The signal was normalized using Iridium 193 as internal standard. The usual observed precision (CV, <10 %) and accuracy (85-115%) from our routine laboratory method were monitored using independent whole blood internal quality controls (Recipe Chemicals^TM^, München, Germany, levels 35, 90 and 250 µg.L^−1^) and by participation in the External Quality Assessment Schemes organized by the European Network of the Occupational and Environmental Laboratory Medicine. Comparability of background signal due to filter paper with reactive blank was assessed by digesting an equivalent area of paper free of blood.

### Data analysis

The threshold for lead poisoning used in this study was 100 μg.L^−1^, which is the previous pre-2015 level selected by the WHO to define childhood lead poisoning (5). We did not use the threshold of 50 μg.L^−1^ because it refers to childhood lead poisoning, and our population did not include children (5). We considered a blood lead level (BLL) above 1000 μg.L^−1^ without signs of encephalopathy to be implausible and thus an outlier. Indeed, asymptomatic BLLs above 1000 μg.L^−1^ appear to be rare in the literature, even in adults [12–14].

The variable “occupation”, i.e. the activity on the gold mine, was divided into two categories according to the contact each activity has with the mud (support activity versus direct mining activity, i.e with contact with mud). The variable “income” was categorized based on a comparison with the minimum wage in Brazil for the previous month: *high* (more than four times the minimum wage), *low* (less than the minimum wage), and *moderate* (between these two ranges). According to previous studies, gold mining sites were geographically grouped into three major regions based on population flows and proximity [15,16]. When the site was unknown, the place of inclusion in the study was used as a proxy for the gold-mining region (18/526, 3.4% of participants). Since the half-life in blood and soft tissues is 30 days, it was assumed that the likely site of contamination was the gold mining site where the miner had been in the previous 30 days [17]. The variable “disposal of used batteries” distinguishes between those who take waste out of the forest and those who leave it on the ground in the forest. “Water treatment” distinguishes between those who treat water taken from wells or rivers in the forest (boiling, decanting, hydrochloric acid) and those who do not.

Data were described using frequencies and percentages for qualitative variables, and medians and interquartile ranges for quantitative variables. Blood lead level (BLL) was summarized using geometric mean and associated 95%CIs. Quantitative variables were categorized according to their median. Associations between variables of interest and BLL were explored with 1) single logistic regression models using BLL as a categorical variable (<100 vs. >100 µg.L^−1^), and 2) single linear regression models using log transformed BLL. Variables associated with a p-value <0.20 were then included in mutiple regression models. When two variables showed strong collinearity (e.g. age and amount of time spent in gold mining), only one was kept in the regression model. The two final multiple regression models (logistic and linear) were selected using the Akaike information criterion (AIC). Odds ratios and BLL Ratios were estimated by exponentiating the coefficients of the logistic and linear regression models, respectively. The geographic location of the supposed lead intoxication was mapped using R software version 4.2.2 (2022-10-31). The main libraries used were: tidyverse version 2.0.0, GEOS version 3.10.2, GDAL version 3.4.2, PROJ version 8.2.1 and the thresholds of 100 and 200 μg.L^−1^.

### Ethical aspects

A written consent form was collected after information in the participant’s language on the primary objective of the study related to malaria and secondary objectives related to heavy metal poisoning.

The study obtained authorization from the Surinamese Ministry of Public Health (N°CMWO005) and from the Brazilian National Research Ethics Commission (N°5.507.241) as part of the Curema project. The authorization of importation of human biological samples was obtained (n°IE-2022-2169) and the biological collection declared to the French Ministry of Education and Research. The database was anonymized and registered according to the General Data Protection Regulation (GDPR). This study was conducted in accordance with the principles set forth in the Declaration of Helsinki.

### Funding

The study was funded by European Interreg Amazon Cooperation Program (N° Presage 8754) and the French Guiana Regional Health Agency. Funding sources did not have any role in the collection and analysis of the data, and in the decision to submit the paper for publication.

### Patient and public involvement

The study population was involved in the development of the questionnaire (with the definition of meaningful questions and appropriate wording), and pre-testing of the questionnaire (comprehension, acceptability, duration). Recruitment of participants was done by opportunistic meeting and then by snowball sampling, thus involving the participants themselves. The questionnaire was administered by a health mediator, who is a pair issued from the target community.

## Results

### Study population

Between September and December 2022, 539 persons were included in the cross-sectional survey. Four subjects were withdrawn for lack of identification on the blotters so 526 participants are included in the present survey on lead poisoning (Fig. 1). The median age was 38 years [Interquantile Range (IQR)=30-48]. Men represented 73.5% of the sample (387/526). The median length of time spent in mining was 8 years (IQR=2-16).

**Figure 1:**
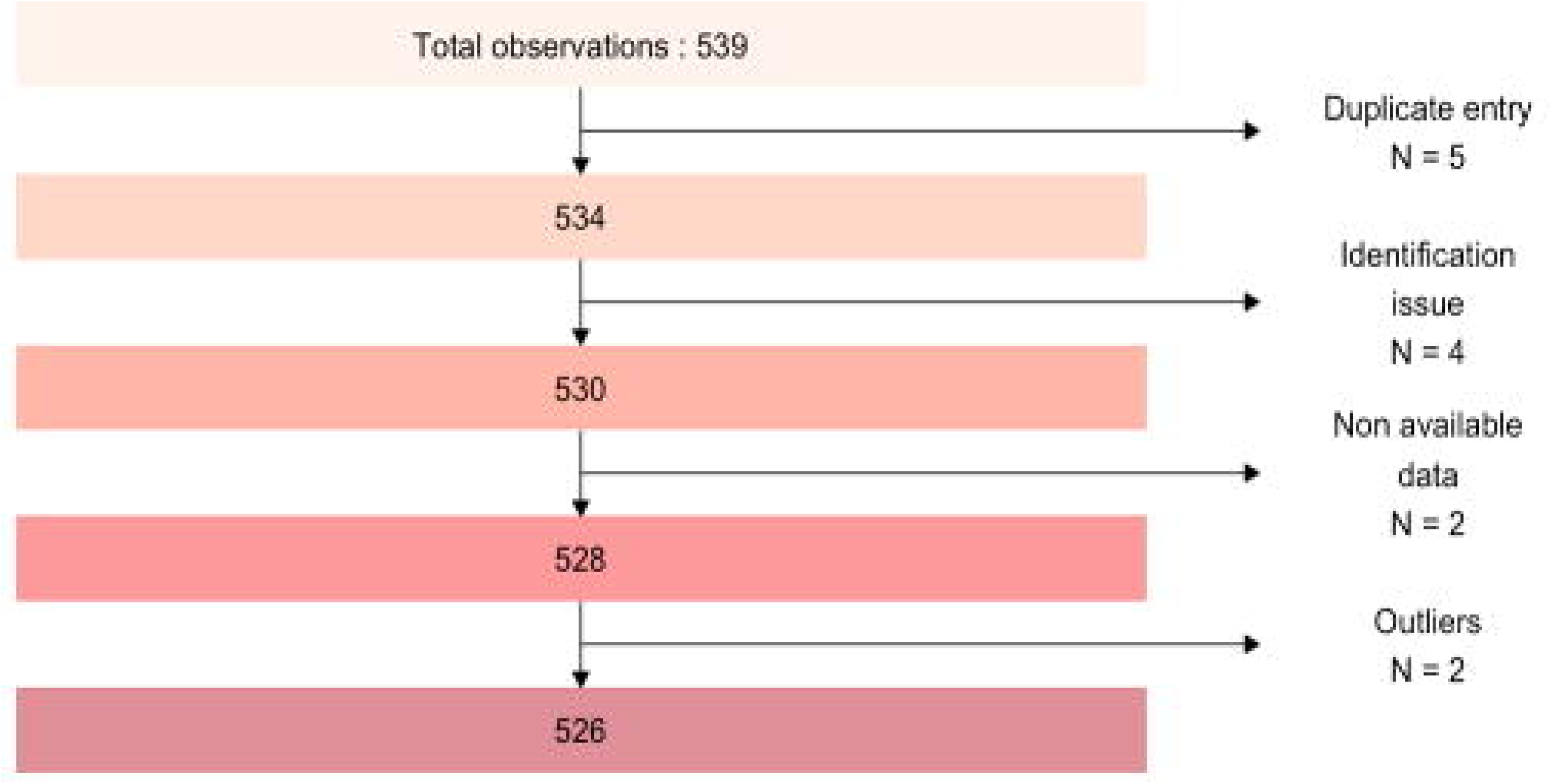
Flow chart of the persons working on the gold mining sites in FG included in the survey, FG, 2022.

### Prevalence of lead poisoning

The prevalence of lead poisoning using the 100 μg.L^−1^ threshold was 44.7 % [40.4-49.0] (Table 1). The median BLL in the study population was 93.6 μg.L^−1^ [IQR: 49.7-152.9], with a maximum of 828.72 μg.L^−1^, and the geometric mean was 95.2 μg.L^−1^ [95%CI: 89.0, 101.8].

**Table 1:** Prevalence of lead poisoning by BLL threshold among people working on gold mining sites, FG, 2022 (N=526)

| Threshold | Prevalence | 95%CI |
| --- | --- | --- |
| 50 $\mu\text{g.L}^{-1}$ | 74.7 % | 70.8-78.4 |
| 100 $\mu\text{g.L}^{-1}$ | 44.7 % | 40.4-49.0 |
| 200 $\mu\text{g.L}^{-1}$ | 15.2 % | 12.3-18.6 |

### Spatial distribution

The likely location of lead contamination shows the difference in repartition in the three identified major mining regions (Fig. 2 and tables 2 and 3).

**Figure 2:**
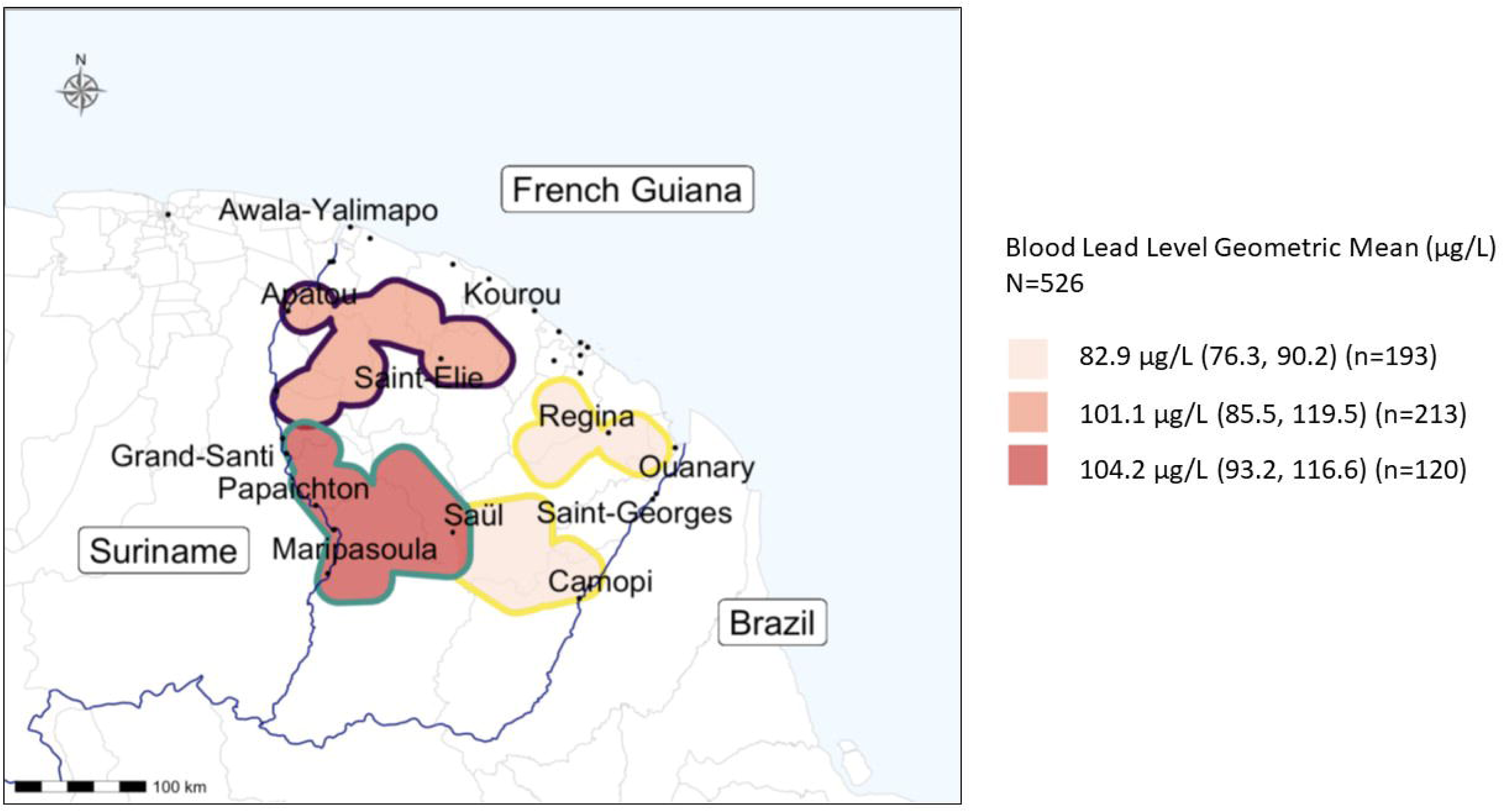
Blood Lead Level geometric mean based on gold-mining regions, FG, 2022.

**Table 2:** Single and multiple logistic regression of factors associated with BLL above 100 μg.L-1 in a sample of individuals working on gold mining sites, FG, 2022 (N=526)

| | N (%) /<br>Median<br>(IQR) | BLL* > 100 $\mu\text{g/L}$<br>% (95%CI) | Single regression<br>OR** (95%CI) | p-value | Multiple<br>regression<br>Adjusted OR<br>(95%CI) |
| --- | --- | --- | --- | --- | --- |
| Sex (n=526) |  |  |  | 0.1 |  |
| Female | 142 (27.0) | 38.7 (30.7, 47.3) | 1 |  |  |
| Male | 384 (73.0) | 46.9 (41.8, 52.0) | 1.40 (0.94, 2.07) |  |  |
| **Age category (n=526)** |  |  |  | 0.02 |  |
| 18-30 yo | 138 (26.2) | 41.3 (33.0, 50.0) | 1 |  |  |
| 31-38 yo | 132 (25.1) | 35.6 (27.5, 44.4) | 0.79 (0.48, 1.28) |  |  |
| 39-48 yo | 129 (24.5) | 50.4 (41.5, 59.3) | 1.44 (0.89, 2.35) |  |  |
| 49-71 yo | 127 (24.1) | 52.0 (42.9, 60.9) | 1.54 (0.95, 2.51) |  |  |
| **Age (10 years) (n=526)** | 3.8 (3.0, 4.8) |  | 1.21 (1.05, 1.40) | 0.008 |  |
| **Weight (n=523)** |  |  |  | 0.9 |  |
| 43-60 kg | 134 (25.6) | 44.8 (36.2, 53.6) | 1 |  |  |
| 61-66 kg | 128 (24.5) | 47.7 (38.8, 56.7) | 1.12 (0.69, 1.83) |  |  |
| 67-74 kg | 137 (26.2) | 43.8 (35.3, 52.5) | 0.96 (0.59, 1.55) |  |  |
| 75-101 kg | 124 (23.7) | 43.5 (34.7, 52.7) | 0.95 (0.58, 1.56) |  |  |
| **Time spent in gold mining, category (n=524)** |  |  |  | 0.02 |  |
| < 8 years | 269 (51.3) | 39.8 (33.9, 45.9) | 1 |  |  |
| > 8 years | 255 (48.7) | 50.2 (43.9, 56.5) | 1.53 (1.08, 2.16) |  |  |
| **Time spent in gold mining (10 years) (n=524)** | 0.8 (0.2, 1.6) |  | 1.17 (0.99, 1.37) | 0.06 | 1.31 (1.09, 1.58) |
| **Time not spent in gold mining (10 years) (n=524)** | 2.7 (2.1, 3.5) |  | 1.10 (0.94, 1.30) | 0.2 | 1.22 (1.01, 1.47) |
| **Time spent on the last mining site (n=518)** |  |  |  | 0.6 |  |
| < 8 months | 269 (51.9) | 43.1 (37.1, 49.3) | 1 |  |  |
| > 8 months | 249 (48.1) | 45.8 (39.5, 52.2) | 1.11 (0.79, 1.58) |  |  |
| **Income (n=498)** |  |  |  | 0.2 |  |
| Low | 92 (18.5) | 52.2 (41.5, 62.7) | 1 |  |  |
| Moderate | 344 (69.1) | 42.4 (37.2, 47.9) | 0.68 (0.43, 1.07) |  |  |
| High | 62 (12.4) | 45.2 (32.5, 58.3) | 0.75 (0.39, 1.44) |  |  |
| **Mode of gold extraction (n=430)** |  |  |  | 0.5 |  |
| Alluvial | 361 (84.0) | 46.8 (41.6, 52.1) | 1 |  |  |
| Pit or other | 69 (16.0) | 52.2 (39.8, 64.4) | 1.24 (0.74, 2.08) |  |  |
| **Mobility (n=506)** |  |  |  | 0.06 |  |
| Moderate | 301 (59.5) | 48.5 (42.7, 54.3) | 1 |  |  |
| Important | 205 (40.5) | 39.5 (32.8, 46.6) | 0.69 (0.48, 0.99) |  |  |
| **Type of occupation (n=506)** |  |  |  | 0.004 |  |
| Support activity | 201 (39.7) | 36.8 (30.1, 43.9) | 1 |  | 1 |
| Direct mining activity | 305 (60.3) | 50.2 (44.4, 55.9) | 1.73 (1.20, 2.49) |  | 1.67 (1.13, 2.48) |
| **Gold mining region (n=526)** |  |  |  | 0.03 |  |
| East | 193 (36.7) | 38.3 (31.5, 45.6) | 1 |  | 1 |
| South-Western | 213 (40.5) | 51.2 (44.3, 58.1) | 1.69 (1.14, 2.51) |  | 2.09 (1.34, 3.27) |
| North-Western | 120 (22.8) | 43.3 (34.3, 52.7) | 1.23 (0.77, 1.95) |  | 1.58 (0.96, 2.63) |
| **Water treatment (n=516)** |  |  |  | 0.02 |  |
| No | 353 (68.4) | 41.1 (35.9, 46.4) | 1 |  |  |
| Yes | 163 (31.6) | 52.1 (44.2, 60.0) | 1.56 (1.08, 2.27) |  |  |
| **Sanitation (n=526)** |  |  |  | 0.1 |  |
| Latrines | 25 (4.8) | 28.0 (12.1, 49.4) | 1 |  |  |
| No latrines | 501 (95.2) | 45.5 (41.1, 50.0) | 2.15 (0.92, 5.61) |  |  |
| **Couac consumption (n=526)** |  |  |  | 0.4 |  |
| Never | 38 (7.2) | 55.3 (38.3, 71.4) | 1 |  |  |
| Casual | 174 (33.1) | 43.7 (36.2, 51.4) | 0.63 (0.31, 1.27) |  |  |
| Daily | 314 (59.7) | 43.9 (38.4, 49.6) | 0.63 (0.32, 1.25) |  |  |
| **Fishing nets (n=526)** |  |  |  | 0.06 |  |
| No fishing | 350 (66.5) | 42.3 (37.1, 47.7) | 1 |  |  |
| No lead weighted nets | 93 (17.7) | 43.0 (32.8, 53.7) | 1.03 (0.65, 1.63) |  |  |
| Lead weighted nets | 83 (15.8) | 56.6 (45.3, 67.5) | 1.78 (1.10, 2.90) |  |  |
| **Hunting practice (n=525)** |  |  |  | 0.008 |  |
| No | 439 (83.6) | 41.9 (37.3, 46.7) | 1 |  |  |
| Yes | 86 (16.4) | 58.1 (47.0, 68.7) | 1.92 (1.21, 3.09) |  |  |
| **Game consumption (n=516)** |  |  |  | 0.06 |  |
| No | 175 (33.9) | 38.3 (31.1, 45.9) | 1 |  | 1 |
| Yes | 341 (66.1) | 47.5 (42.1, 53.0) | 1.46 (1.01, 2.12) |  | 1.58 (1.06, 2.36) |
| **Lead shots ingestion (n=515)** |  |  |  | 0.2 |  |
| No | 319 (61.9) | 42.0 (36.5, 47.6) | 1 |  |  |
| Yes | 196 (38.1) | 48.5 (41.3, 55.7) | 1.30 (0.91, 1.86) |  |  |
| **Disposal of used batteries (n=443)** |  |  |  | 0.9 |  |
| Left on site | 125 (28.2) | 42.4 (33.6, 51.6) | 1 |  |  |
| Else | 318 (71.8) | 43.4 (37.9, 49.0) | 1.04 (0.69, 1.59) |  |  |

\* BLL = Blood Lead Level
\*\* Odds-Ratio

**Table 3:**
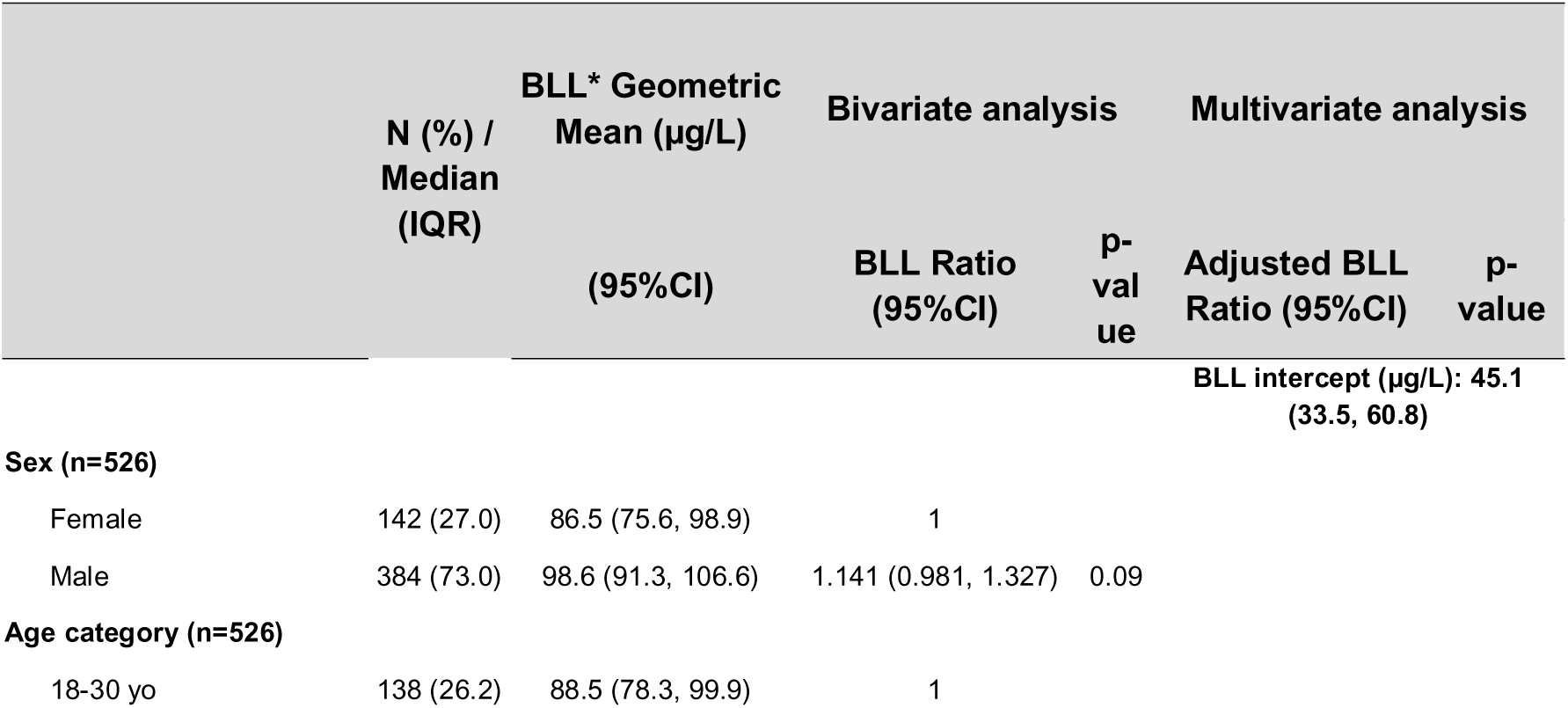
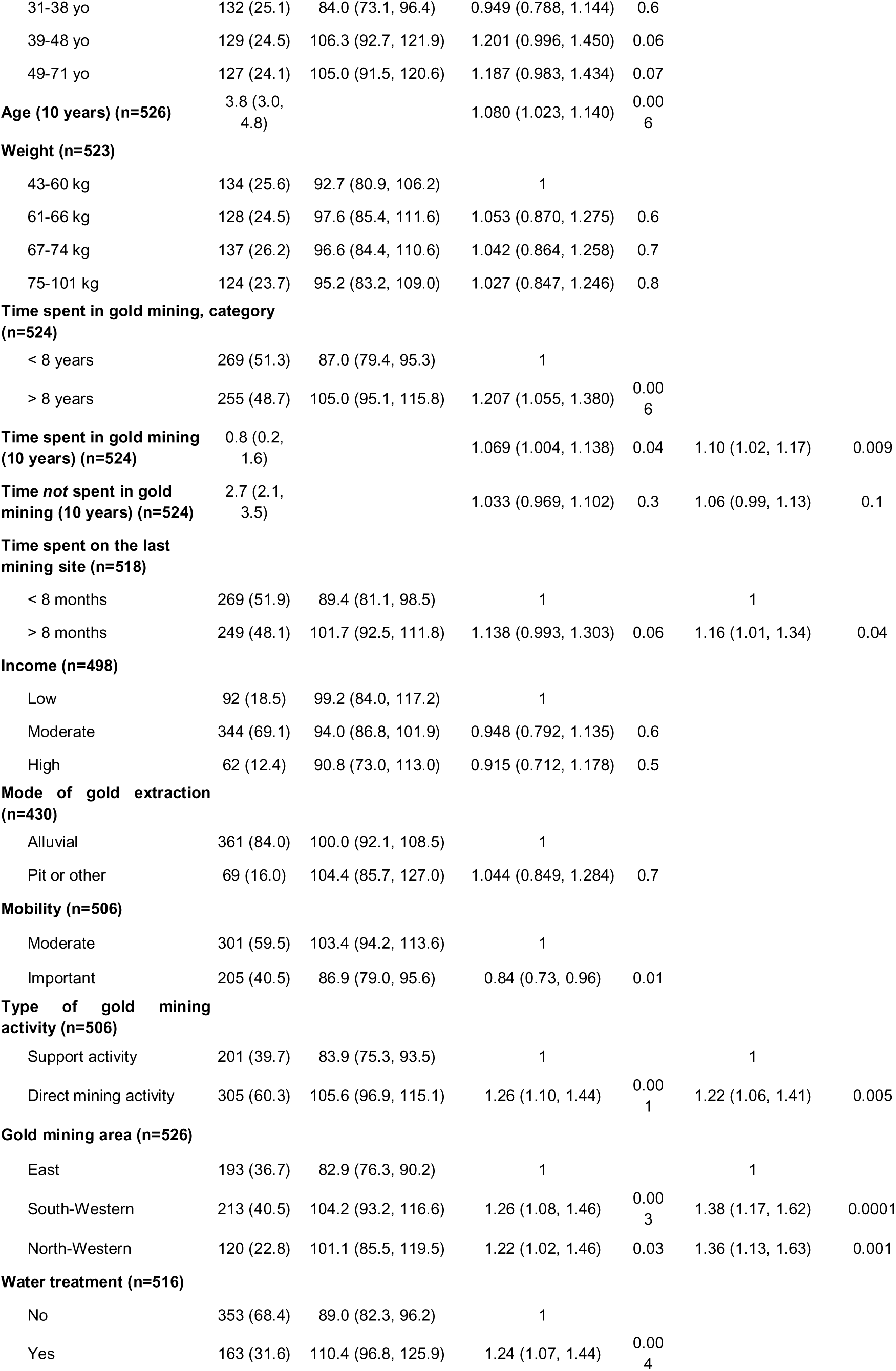
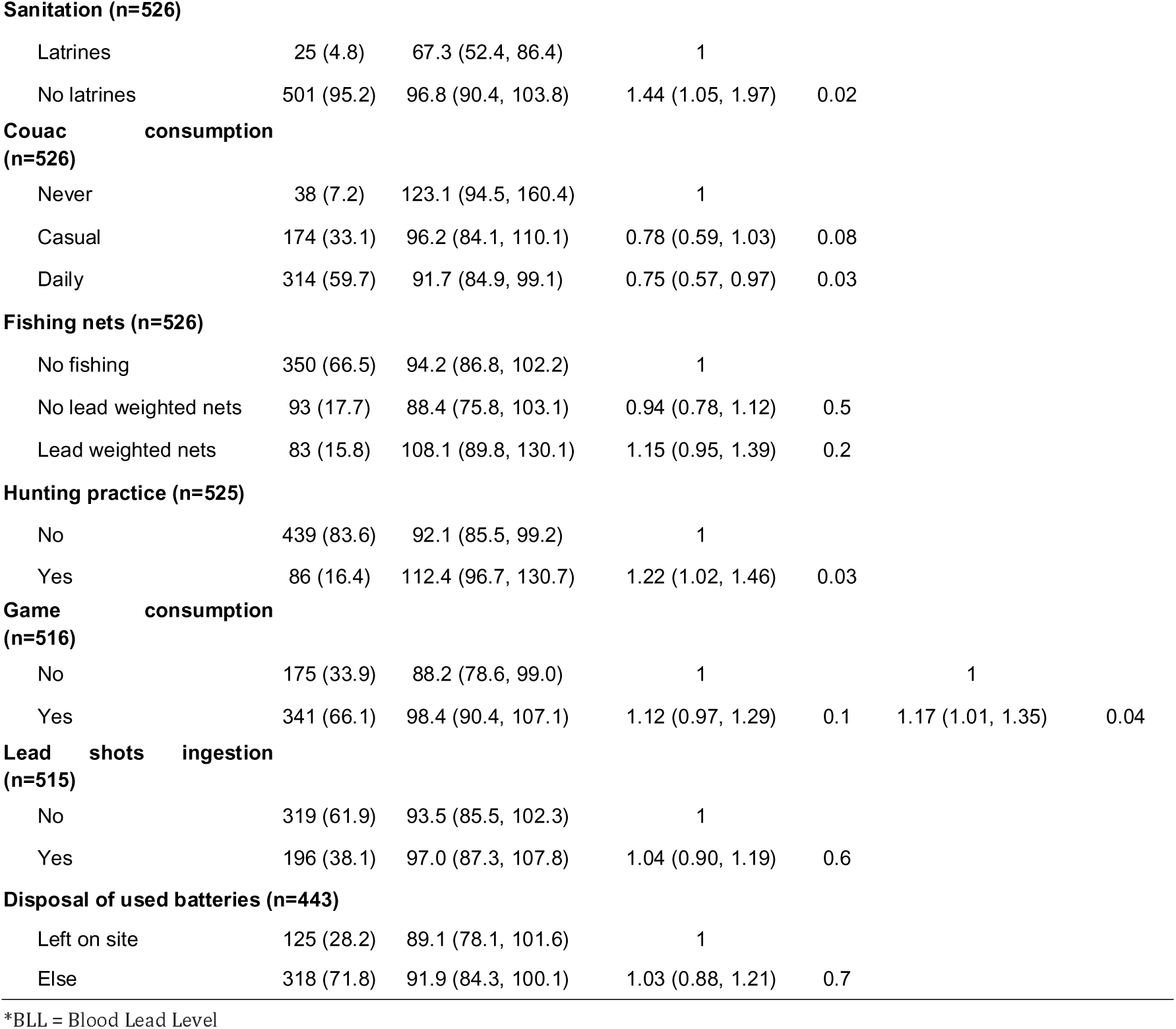
Single and mutiple linear regression of factors associated with log-BLL in a sample of individuals working on gold mining sites, FG, 2022 (N=526)

|  | N (%) /<br>Median<br>(IQR) | BLL* Geometric<br>Mean (µg/L) | Bivariate analysis |  | Multivariate analysis |  |
| --- | --- | --- | --- | --- | --- | --- |
|  |  | (95%CI) | BLL Ratio<br>(95%CI) | p-<br>val<br>ue | Adjusted BLL<br>Ratio (95%CI) | p-value |
| BLL intercept (µg/L): 45.1<br>(33.5, 60.8) |  |  |  |  |  |  |

**Sex (n=526)**
|  |  |  |  |  |
| --- | --- | --- | --- | --- |
| Female | 142 (27.0) | 86.5 (75.6, 98.9) | 1 |  |
| Male | 384 (73.0) | 98.6 (91.3, 106.6) | 1.141 (0.981, 1.327) | 0.09 |
| **Age category (n=526)** |  |  |  |  |  |  |
| 18-30 yo | 138 (26.2) | 88.5 (78.3, 99.9) | 1 |  |  |  |
| 31-38 yo | 132 (25.1) | 84.0 (73.1, 96.4) | 0.949 (0.788, 1.144) | 0.6 |  |  |
| 39-48 yo | 129 (24.5) | 106.3 (92.7, 121.9) | 1.201 (0.996, 1.450) | 0.06 |  |  |
| 49-71 yo | 127 (24.1) | 105.0 (91.5, 120.6) | 1.187 (0.983, 1.434) | 0.07 |  |  |
| **Age (10 years) (n=526)** | 3.8 (3.0, 4.8) |  | 1.080 (1.023, 1.140) | 0.006 |  |  |
| **Weight (n=523)** |  |  |  |  |  |  |
| 43-60 kg | 134 (25.6) | 92.7 (80.9, 106.2) | 1 |  |  |  |
| 61-66 kg | 128 (24.5) | 97.6 (85.4, 111.6) | 1.053 (0.870, 1.275) | 0.6 |  |  |
| 67-74 kg | 137 (26.2) | 96.6 (84.4, 110.6) | 1.042 (0.864, 1.258) | 0.7 |  |  |
| 75-101 kg | 124 (23.7) | 95.2 (83.2, 109.0) | 1.027 (0.847, 1.246) | 0.8 |  |  |
| **Time spent in gold mining, category (n=524)** |  |  |  |  |  |  |
| < 8 years | 269 (51.3) | 87.0 (79.4, 95.3) | 1 |  |  |  |
| > 8 years | 255 (48.7) | 105.0 (95.1, 115.8) | 1.207 (1.055, 1.380) | 0.006 |  |  |
| **Time spent in gold mining (10 years) (n=524)** | 0.8 (0.2, 1.6) |  | 1.069 (1.004, 1.138) | 0.04 | 1.10 (1.02, 1.17) | 0.009 |
| **Time not spent in gold mining (10 years) (n=524)** | 2.7 (2.1, 3.5) |  | 1.033 (0.969, 1.102) | 0.3 | 1.06 (0.99, 1.13) | 0.1 |
| **Time spent on the last mining site (n=518)** |  |  |  |  |  |  |
| < 8 months | 269 (51.9) | 89.4 (81.1, 98.5) | 1 |  | 1 |  |
| > 8 months | 249 (48.1) | 101.7 (92.5, 111.8) | 1.138 (0.993, 1.303) | 0.06 | 1.16 (1.01, 1.34) | 0.04 |
| **Income (n=498)** |  |  |  |  |  |  |
| Low | 92 (18.5) | 99.2 (84.0, 117.2) | 1 |  |  |  |
| Moderate | 344 (69.1) | 94.0 (86.8, 101.9) | 0.948 (0.792, 1.135) | 0.6 |  |  |
| High | 62 (12.4) | 90.8 (73.0, 113.0) | 0.915 (0.712, 1.178) | 0.5 |  |  |
| **Mode of gold extraction (n=430)** |  |  |  |  |  |  |
| Alluvial | 361 (84.0) | 100.0 (92.1, 108.5) | 1 |  |  |  |
| Pit or other | 69 (16.0) | 104.4 (85.7, 127.0) | 1.044 (0.849, 1.284) | 0.7 |  |  |
| **Mobility (n=506)** |  |  |  |  |  |  |
| Moderate | 301 (59.5) | 103.4 (94.2, 113.6) | 1 |  |  |  |
| Important | 205 (40.5) | 86.9 (79.0, 95.6) | 0.84 (0.73, 0.96) | 0.01 |  |  |
| **Type of gold mining activity (n=506)** |  |  |  |  |  |  |
| Support activity | 201 (39.7) | 83.9 (75.3, 93.5) | 1 |  | 1 |  |
| Direct mining activity | 305 (60.3) | 105.6 (96.9, 115.1) | 1.26 (1.10, 1.44) | 0.001 | 1.22 (1.06, 1.41) | 0.005 |
| **Gold mining area (n=526)** |  |  |  |  |  |  |
| East | 193 (36.7) | 82.9 (76.3, 90.2) | 1 |  | 1 |  |
| South-Western | 213 (40.5) | 104.2 (93.2, 116.6) | 1.26 (1.08, 1.46) | 0.003 | 1.38 (1.17, 1.62) | 0.0001 |
| North-Western | 120 (22.8) | 101.1 (85.5, 119.5) | 1.22 (1.02, 1.46) | 0.03 | 1.36 (1.13, 1.63) | 0.001 |
| **Water treatment (n=516)** |  |  |  |  |  |  |
| No | 353 (68.4) | 89.0 (82.3, 96.2) | 1 |  |  |  |
| Yes | 163 (31.6) | 110.4 (96.8, 125.9) | 1.24 (1.07, 1.44) | 0.004 |  |  |
| **Sanitation (n=526)** |  |  |  |  |  |  |
| Latrines | 25 (4.8) | 67.3 (52.4, 86.4) | 1 |  |  |  |
| No latrines | 501 (95.2) | 96.8 (90.4, 103.8) | 1.44 (1.05, 1.97) | 0.02 |  |  |
| **Couac consumption (n=526)** |  |  |  |  |  |  |
| Never | 38 (7.2) | 123.1 (94.5, 160.4) | 1 |  |  |  |
| Casual | 174 (33.1) | 96.2 (84.1, 110.1) | 0.78 (0.59, 1.03) | 0.08 |  |  |
| Daily | 314 (59.7) | 91.7 (84.9, 99.1) | 0.75 (0.57, 0.97) | 0.03 |  |  |
| **Fishing nets (n=526)** |  |  |  |  |  |  |
| No fishing | 350 (66.5) | 94.2 (86.8, 102.2) | 1 |  |  |  |
| No lead weighted nets | 93 (17.7) | 88.4 (75.8, 103.1) | 0.94 (0.78, 1.12) | 0.5 |  |  |
| Lead weighted nets | 83 (15.8) | 108.1 (89.8, 130.1) | 1.15 (0.95, 1.39) | 0.2 |  |  |
| **Hunting practice (n=525)** |  |  |  |  |  |  |
| No | 439 (83.6) | 92.1 (85.5, 99.2) | 1 |  |  |  |
| Yes | 86 (16.4) | 112.4 (96.7, 130.7) | 1.22 (1.02, 1.46) | 0.03 |  |  |
| **Game consumption (n=516)** |  |  |  |  |  |  |
| No | 175 (33.9) | 88.2 (78.6, 99.0) | 1 |  | 1 |  |
| Yes | 341 (66.1) | 98.4 (90.4, 107.1) | 1.12 (0.97, 1.29) | 0.1 | 1.17 (1.01, 1.35) | 0.04 |
| **Lead shots ingestion (n=515)** |  |  |  |  |  |  |
| No | 319 (61.9) | 93.5 (85.5, 102.3) | 1 |  |  |  |
| Yes | 196 (38.1) | 97.0 (87.3, 107.8) | 1.04 (0.90, 1.19) | 0.6 |  |  |
| **Disposal of used batteries (n=443)** |  |  |  |  |  |  |
| Left on site | 125 (28.2) | 89.1 (78.1, 101.6) | 1 |  |  |  |
| Else | 318 (71.8) | 91.9 (84.3, 100.1) | 1.03 (0.88, 1.21) | 0.7 |  |  |

\*BLL = Blood Lead Level

### Bi and Multivariate analyses

Table 2 shows the results of the single and mutiple regression models. The factors associated with a BLL over 100 μg.L^−1^ were: amount of time spent in gold mining (OR=1.31 [1.09-1.58], i.e. among participants, the odds of having a BLL above 100 µg.L^−1^ increased by 1.31 for each decade spent in gold mining), a mining occupation in direct contact with mud (OR=1.67 [1.13-2.48]), working in the Southwestern region (OR=2.09 [1.34-3.27]) and consuming game (OR=1.58 [1.06-2.36]).

The linear model assessed the variables associated with the log of the BLL (Table 3). The BLL intercept was 45.1 μg.L^−1^ [33.5-60.8].BLL was 1.22 times higher among participants with an occupation with direct mud exposure, compared to participants without mud exposure. For each decade spent in gold mining, BLL increased by 10% (BLL Ratio=1.10).

### Potential Health Impact

One woman was pregnant at the time of inclusion and had a BLL of 59.9 μg.L^−1^. Four other women did not know if they were pregnant and had BLLs of 37.2, 96.7, 110.5, 115.3 μg.L^−1^. Medical examination did not reveal any neurological disorders. Among the study population, 18.9% [15.7-22.5] had a blood pressure above 140/90 mmHg and 5.3% [3.6-7.6] above 160/100. Hypertension was not associated with BLL, either quantitatively (p=0.257) or in terms of systolic blood pressure above 160 mmHg (p=0.374), with no difference between genders. One third of the study population (39.5%) declared that digestive disorders were among the top three health problems at the sites.

## Discussion

This study shows that, using the 100 μg.L^−1^ threshold, the prevalence of lead poisoning among gold miners was staggeringly high — 44.7% [40.4-49.0]. The main factors associated with lead poisoning were the time spent in gold mining activity, occupation with direct exposure to mud compared to support activities, the region of gold mining and game consumption. The risk factors presumed in the French Guianese population (such as couac consumption) were not found. Taken together, these results argue for environmental contamination.

### A staggering level of lead poisoning among gold miners

The prevalence of lead poisoning in the study population (44.7%) was significantly higher than in other populations in FG. Thus, the Guyaplomb study estimated the prevalence of lead poisoning among Guyanese children aged 1 to 6 years in 2015 to be at 20.1%,using the pediatric threshold of 50 μg.L^−1^ [7]. Another study, among pregnant women in the western part of FG, found that 25.8% and 5.1% of these women had lead levels above 50 μg.L^−1^ and 100 μg.L^−1^ respectively [18]. In neighboring Suriname, the prevalence of lead levels above 35 µg.L^−1^ in pregnant women was 21% [19]. In Brazil, a study of 448 individuals from 12 indigenous communities in the nearby state of Pará reported, a prevalence of 79.0% and 35.8% in their sample of men and women, respectively at the threshold of 100 μg.L^−1^ [20]. In comparison, in mainland France, using the threshold of 100 μg.L^−1^, the prevalence of lead poisoning was estimated to be 1.7 % among adults aged 18 to 74 years [21]. This emphasizes that lead poisoning is a major public health threat in FG, both in the French Guianese population and among gold miners [22].

### Arguments for environmental exposure

In mainland France, the main sources of lead exposure are old lead paint flakes, pipes containing lead, or localized exposure to soil contaminated by industrial or mining activities [1,3,23]. These factors cannot be extrapolated to the Amazon region. The factors cited in French Guiana (couac consumption or accidental ingestion of lead shots [8,18,24]) have also been reported in other Amazonian regions: consumption of wild game in Peru, or cassava flour, which is equivalent of couac, in Brazil [20,25,26]. The bioaccumulation of natural lead in cassava tubers is a hypothesis currently being evaluated in the Amerindian population [24]. The artisanal transformation of cassava into flour (farinha), which requires heating in large metal tool has been associated with higher levels of lead [20]. However, in our population, no association was found with the consumption of *couac*, whether locally or industrially produced. While the reported ingestion of lead shot was not significant, a statistical association between BLL and game consumption was observed. This could be due to the diffusion of lead into the meat following hunting with lead shots, even if the shots were removed before consumption [27,28]. In France lead shots are regulated and banned in all wetlands and for waterbird hunting [29]. In French Guiana it is still commonly used. However lead poisoning was associated with an activity directly related to the mud, the mining region and with the amount of time spent in gold mining. In neighboring Suriname, there was also spatial heterogeneity, with a higher lead exposure among pregnant women living in the interior of the country [19]. This raises the hypothesis that contamination may originate from naturally occuring telluric lead. Indeed, gold and lead can be intertwined in the rocks and lead contamination in gold mining settlements has been demonstrated in several settings [30–33]. The route of contamination remains to be investigated. Contamination could be digestive, e.g. related to drinking water, for example, as gold miners do not have access to drinking water [10], but it could also be due to inhalation of particles in suspension. Geological surveys are underway throughout FG to study the composition of the soils and mayprovide more information on the relationship between gold and lead on this territory. The impact of gold mining on the release of lead downstream of mines into the environment also needs to be studied.

### Challenges for individual and public health

The study population only concerned adults, with few pregnant or presumed pregnant women. However, 75% (104/139) of the women were of childbearing age and during pregnancy, lead stored in the bones of an exposed mother is released into the bloodstream, thereby contaminating the fetus [34]. The consequences for the fetus, particularly the neurodevelopmental and neurobehavioral impacts of lead are well described [35]. Some children occasionally come to gold mines with their parents, but this study did not assess whether they are exposed to lead in the same way as adults. Health professionals treating children who have spent time in gold mines should measure their BLL.

Adults who work in gold mines often report abdominal pain attributed to digestive parasitosis but could also be a manifestation of lead poisoning. This study did not demonstrate an association between high blood pressure and lead poisoning as reported in the literature and did not allow evaluation of an association with anemia or cardiopathies [36–38]. This would require further evaluation.

### Limitations

Association with a presumed exposure zone is subject to bias because of the high mobility of gold miners and the chronic nature of the contamination. Because the data were collected on a declarative basis, there may be recall or desired response bias.

## Conclusion

This study confirms that lead poisoning is a major issue in FG and provides food for thought about the risk factors for lead poisoning, which are still poorly understood in this region. Indeed, the usual hypothetical risk factors in French Guiana, except game consumption, were not associated with lead levels. By contrast, our observations converged to suggest an environmental source. Subsequent multidisciplinary studies involving toxicology, geology, epidemiology and sociology will make it possible to refine knowledge and propose prevention strategies. A regional network on this subject is being set up by the FG Regional Health Agency.

## Glossary

ASGM: Artisanal and Small-scale Gold Mining
BMIG: Brazilian Minimum Interprofessional Growth Wage (around 260 US dollars in 2023)
BLL: Blood Lead Level
CI95: Confidence Intervalle (95%)
CUREMA: Radical CURE for MAlaria among highly mobile and hard-to-reach populations in the Guiana Shield
HCSP: Haut Conseil de la Santé Publique
ICP-MS: Inductively Coupled Plasma Mass Spectrometry
OR: Odds ratio
WHO: World Health Organization

## Data Availability

All data produced in the present study are available upon reasonable request to the authors

## Acknowledgement

We thank the Biobank Amazonie from Cayenne’s Hospital and all the participants to the survey, as well as Fabrice Quet for his statistical support.

## Author contribution

MD, VK, YL and OM developed the first draft of the study protocol with support from SV, AS and MSM. LP, TB, MD, SV and MSM conducted the survey on the field. OM performed laboratory analyses. VK performed data analysis with support from YL, MD, AA and MN. All authors contributing to revising the manuscript and provided approval of the final version. MD acted as the guarantor.

## Data Availability Statement

Data are available upon reasonable request from the corresponding author.

## Ethical approval

The protocol was approved by the National Ethics Board of Suriname (CMWO (Commissie voor Mensgebonden Wetenschappelijk Onderzoek), N°005/22) and of Brazil (CONEP (Comissão Nacional de Etica em Pesquisa), N° 5.507.241).

## Patient consent

All participants provided an informed and written consent form.

